# Implementing Building-Level SARS-CoV-2 Wastewater Surveillance on a University Campus

**DOI:** 10.1101/2020.12.31.20248843

**Authors:** Cynthia Gibas, Kevin Lambirth, Neha Mittal, Md Ariful Islam Juel, Visva Bharati Barua, Lauren Roppolo Brazell, Keshawn Hinton, Jordan Lontai, Nicholas Stark, Isaiah Young, Cristine Quach, Morgan Russ, Jacob Kauer, Bridgette Nicolosi, Srinivas Akella, Wenwu Tang, Don Chen, Jessica Schlueter, Mariya Munir

## Abstract

The COVID-19 pandemic has been a source of ongoing challenges and presents an increased risk of illness in group environments, including jails, long term care facilities, schools, and, of course, residential college campuses. Early reports that the SARS-CoV-2 virus was detectable in wastewater in advance of confirmed cases sparked widespread interest in wastewater based epidemiology (WBE) as a tool for mitigation of COVID-19 outbreaks. One hypothesis was that wastewater surveillance might provide a cost-effective alternative to other more expensive approaches such as pooled and random testing of groups. In this paper, we report the outcomes of a wastewater surveillance pilot program at the University of North Carolina at Charlotte, a large urban university with a substantial population of students living in on-campus dormitories. Surveillance was conducted at the building level on a thrice-weekly schedule throughout the university’s fall residential semester. In multiple cases, wastewater surveillance enabled identification of asymptomatic COVID-19 cases that were not detected by other components of the campus monitoring program, which also included in-house contact tracing, symptomatic testing, scheduled testing of student athletes, and daily symptom reporting. In the context of all cluster events reported to the University community during the fall semester, wastewater-based testing events resulted in identification of smaller clusters than were reported in other types of cluster events. Wastewater surveillance was able to detect single asymptomatic individuals in dorms with total resident populations of 150-200. While the strategy described was developed for COVID-19, it is likely to be applicable to mitigation of future pandemics in universities and other group-living environments.

## Introduction

Wastewater based epidemiology (WBE) has been a successful approach to address public health issues such as viral [1–3] and bacterial [4] disease outbreaks and substance abuse [5], with many other potential uses proposed [6]. WBE approaches are often used at the community level to monitor overall abundance of a viral or chemical signal and to guide public health interventions. Early in the COVID-19 pandemic, it was hypothesized that wastewater surveillance could be a cost-effective alternative to high-frequency diagnostic testing of populations [7], and could be used to guide more immediate and targeted interventions including testing of affected building populations. Several universities [8–11] piloted this approach as part of their pandemic reopening strategy in Fall 2020, including UNC Charlotte.

UNC Charlotte implemented an institution-wide monitoring program to mitigate COVID-19 outbreaks during the Fall 2020 semester. The university chose a four-pronged approach that included wastewater surveillance, symptomatic testing, daily health checks, and in-house contact tracing. Student athletes are also tested at a mandated frequency. While it is not our purpose in this paper to describe the details of the other components of the response, the information gathered via in-house testing, contact tracing, and surveys does play an important role in how the wastewater surveillance data is used within the institution.

Wastewater surveillance has been implemented on residential buildings only, primarily at the building level, although in a few cases larger buildings have been split into zones, or smaller buildings such as fraternity houses grouped together in small neighborhoods. A positive wastewater sample may trigger a surge testing event, in which all students are asked to stay in place and a testing team is sent to test everyone in the building. However, wastewater information is first considered in conjunction with other evidence. If a wastewater positive occurs in the absence of previously identified infected persons or known close contacts, then the affected student population is required to remain in their dormitory and to participate in surge testing.

During the shortened fall semester, UNC Charlotte’s combined approach has proven successful at sustaining a de-densified population of students in university residence halls. The university has maintained the ability to offer a mix of instruction including in-person courses, without resorting to repeated mass-testing of the entire campus population. Wastewater monitoring has successfully provided early warning of the spread of COVID-19 in individual dormitory buildings, in some cases detecting a single presymptomatic individual in a dormitory population of 150+, and allowing for their isolation prior to further spread of the virus.

In designing the building-level wastewater surveillance program at UNC Charlotte, we identified six key areas in which methodological choices needed to be made: 1) where to sample, 2) how often to sample, 3) what type of sample to collect, 4) what type of sample concentration protocol to use, 5) what kind of RNA extraction protocol to use, and 6) what type of detection method to use. However, the implementation timeline for the program did not allow for leisurely optimization of any of these protocol choices. The wastewater monitoring component of the campus COVID-19 prevention effort was authorized by the university’s Chancellor on July 27, 2020, with in-person classes originally scheduled to start on September 7, 2020. Events on other UNC system campuses, which had begun their semester in mid-August, prompted a further 3-week delay in the start of on-campus instruction at UNC Charlotte. Student move-in to the dormitories occurred on September 28th and 29th, 2020. With approximately eight weeks to purchase equipment and implement the program once funding was secured, we were required to make initial choices about deployment of equipment and protocols based on relatively limited information. This information was often gleaned from preprints, webinars, and informal discussions. The program has been optimized on the fly during fall semester while maintaining a consistent and actionable sampling routine.

## Methods

### Study Area

UNC Charlotte’s on-campus housing comprises 17 dormitories and a “Greek Village” of multiple small buildings (see Figure 1). A few thousand additional students live in large private apartment complexes adjacent to the University. The dormitory capacity at UNC Charlotte is nearly 8000 students, but as a de-densification strategy, this was reduced to under 4000 for Fall semester 2020. One dormitory was selected as the University’s isolation space for active COVID-19 cases, while two Greek Village buildings were designated as quarantine space for suspected exposures.

**Figure 1.**
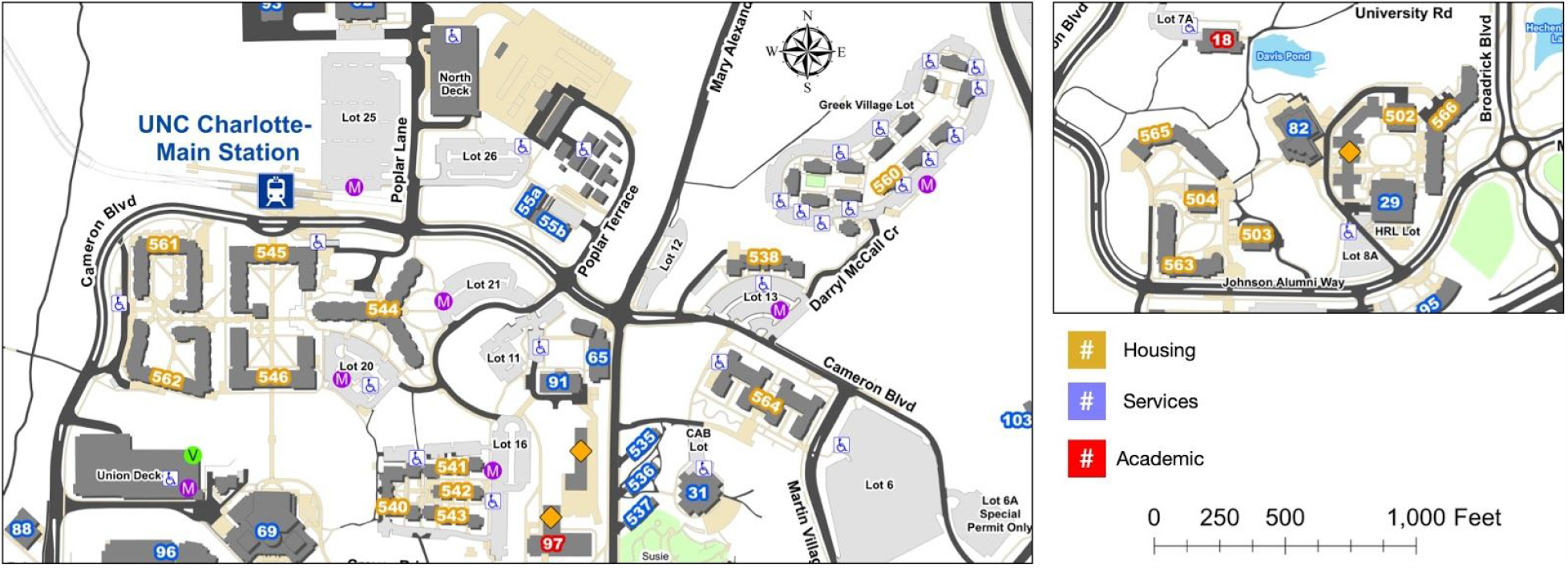
Residential areas of UNC Charlotte Main Campus monitored in this study. Buildings are purposely not identified by name, consistent with University policy regarding reporting of COVID-related data. Left panel: north campus residential areas. Right panel: South Village residential area.

The initial design of the surveillance strategy implemented 19 on-campus sampling points, one in each dormitory, and three manhole sites within the Greek Village. One dormitory building could not be isolated from other wastewater sources. The COVID-19 isolation space could be monitored separately from all but one other building, and served as a type of positive control for the campus. Samples from dormitory sites were collected three times each week during the period of Sep. 28, 2020 – Nov. 23, 2020, the period during which dorm occupancy was at its highest.

### Selection and Optimization of Sampling Sites

Campus wastewater could be readily accessed in two types of locations – inside buildings, directly from building plumbing cleanouts, or at external sites accessed via manholes. At external manhole sites, there was potential that wastewater could be significantly diluted by other building outflows, especially grey water from laundry rooms, showers, and other facilities. Early reports of detection of COVID-19 in wastewater [12–14] were primarily based on settled influents at wastewater treatment plants, and less information was available about appropriate concentration methods for raw sewage from smaller sewersheds or individual buildings. We initially chose to avoid collection of sewage mixed with grey water, to avoid excessive dilution of input material.

Wastewater lines were accessed via standard PVC caps, drilled and fitted with sealed barbed fittings on each side, to which a hand pump or an autosampler could be connected. These access points could be easily installed, and because they used standard plumbing fittings, were straightforward to construct and move. However, we soon discovered that flow at many of these interior building sites was too low to be sampled reliably. We accessed plumbing blueprints for each building to identify the exact wings or rooms in the structure served by that section of plumbing. In several cases, we found that only a small fraction of the building was being sampled.

We then engaged the UNC Charlotte Facilities Management group to construct and install Phase II manhole access points to aggregate wastewater from entire building populations. Autosamplers were anchored to concrete pads adjacent to the manhole sites to prevent theft or tampering, and collection tubes were anchored within the stream of wastewater flow using PVC guides. Despite early concerns about dilution, samples obtained at manhole sites have proven to be easily handled with our standard laboratory protocols and to be sufficiently concentrated to allow for detection of single presymptomatic individuals within a larger building population. Site relocations were largely completed during October and early November, and with the arrival of additional autosampling equipment, we will sample 30 campus residential and neighborhood sites during the spring semester.

### Reporting Cycle and Laboratory Timeline

Using current protocols, the wastewater surveillance laboratory is able to provide a readout of positive wastewater signal for campus sites within 26-30 hours of sample collection. This timeline (Figure 2) has shortened as lab staff have become more familiar with processes, and as the lab has been fully equipped and built out. The laboratory runs three weekly cycles of samples. Each cycle begins with the setup of autosamplers and initiation of a 24 hour composite sampling protocol on Day 1. In the morning of Day 2, samples are collected and returned to the laboratory for processing. The major processing steps include an HA (electronegative) filtration process, which is the most time consuming of the processing protocols, followed by RNA extraction from the collected filter. On the morning of Day 3, qPCR detection is set up and performed, and the laboratory reports to the university administration with a target deadline of 2pm approximately 30 hours following sample collection. This allows sufficient time for consultation with the testing center and contact tracing groups before the end of the business day. University stakeholders can determine that a dormitory showing positive signal will be “locked down” overnight as early as 36 hours after sample collection, and students tested beginning the following morning.

**Figure 2:**
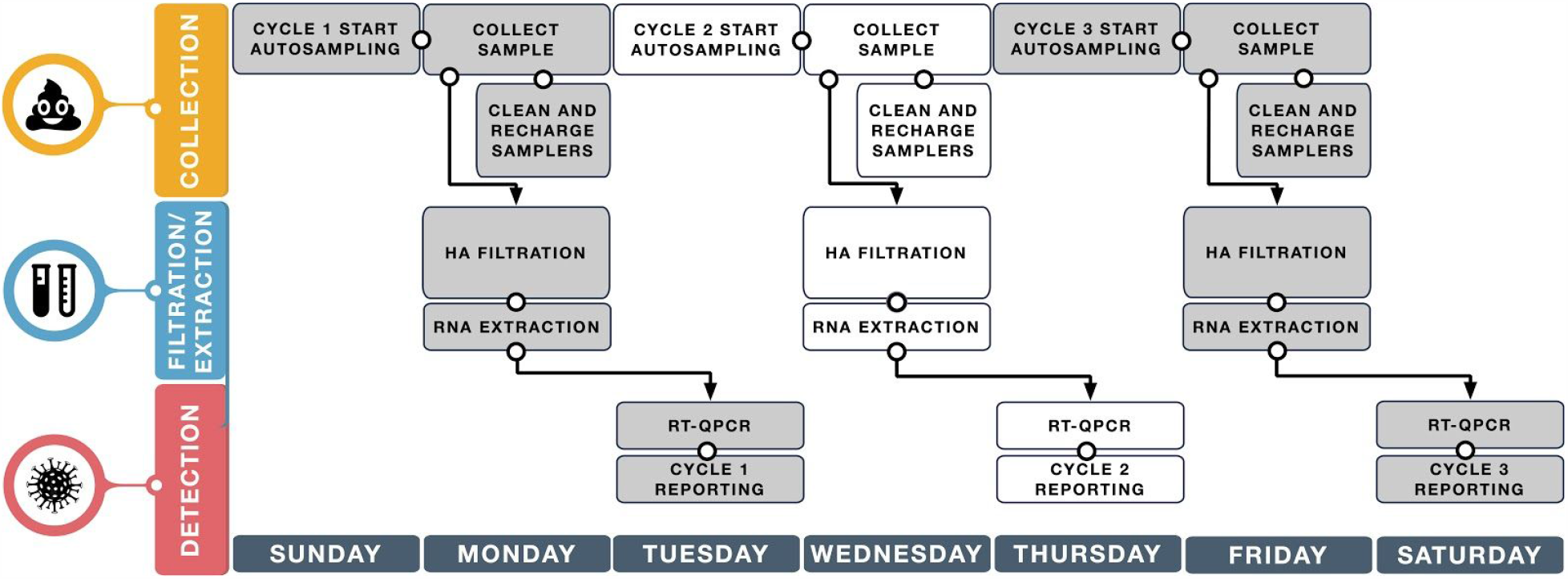
Weekly sample collection and processing timeline for the UNC Charlotte wastewater surveillance project.

### Composite Sample Collection

A variety of sample collection methods are available for building wastewater collection including grab samples (filling a container at one time point), composite samples (manually or mechanically collecting over an extended time window), and Moore swabs [15]. Early studies [16] suggested that while grab and composite sampling produce similar results, composite sample data is ultimately more reliable for modeling purposes, and we chose to pursue that option. Autosamplers used in the UNC Charlotte project include ISCO GLS compact and Hach AS950 portable autosamplers, each loaded with sterilized and acid-washed PET screw-cap carboys in 1-gallon and 2.5-gallon volumes respectively. As many sites are located outdoors, sealed lead-acid AGM batteries are used as power sources for each sampler and recharged daily in between collection events. Vinyl suction tubing of⅜” inner diameter installed with low-flow stainless steel pickup strainers was used for all units at varying lengths of 15-25 feet depending on the depth required of the access points. Setup for each type of installation differed in that manhole locations allowed for direct visual placement of the weighted steel strainer within the flowing wastewater stream, and the additional capability of semi-permanently anchoring the strainer against the flow to maximize collection with 1-inch PVC pipe placed within the manhole. Cleanout pipes were of much smaller diameter, requiring the tapping of removable PVC access caps with barbed coupling fittings or bulkhead fittings to allow tubing attachment and sample flow through the cap. Additionally, the anti-clog pickup holes located on the upper length of the strainer were blocked off with heat shrink tubing to allow suction from the single bottom orifice. This was necessary as cleanout pipes do not contain stagnant water or continuous wastewater flow; therefore the strainer is never submerged and must be placed vertically within the stack, resting the end of the strainer within against the bottom of the pipe (Figure 3).Composite sampling is set to collect over a 24-hour period, with individual 20mL pulls intervaled at 10-30 minutes depending on the installation site type (manhole or cleanout). Cleanouts pulled smaller, more frequent aliquots to increase the probability of a successful grab, whereas manholes present with more constant flow characteristics allowing larger and less frequent pulls of volume.

**Figure 3:**
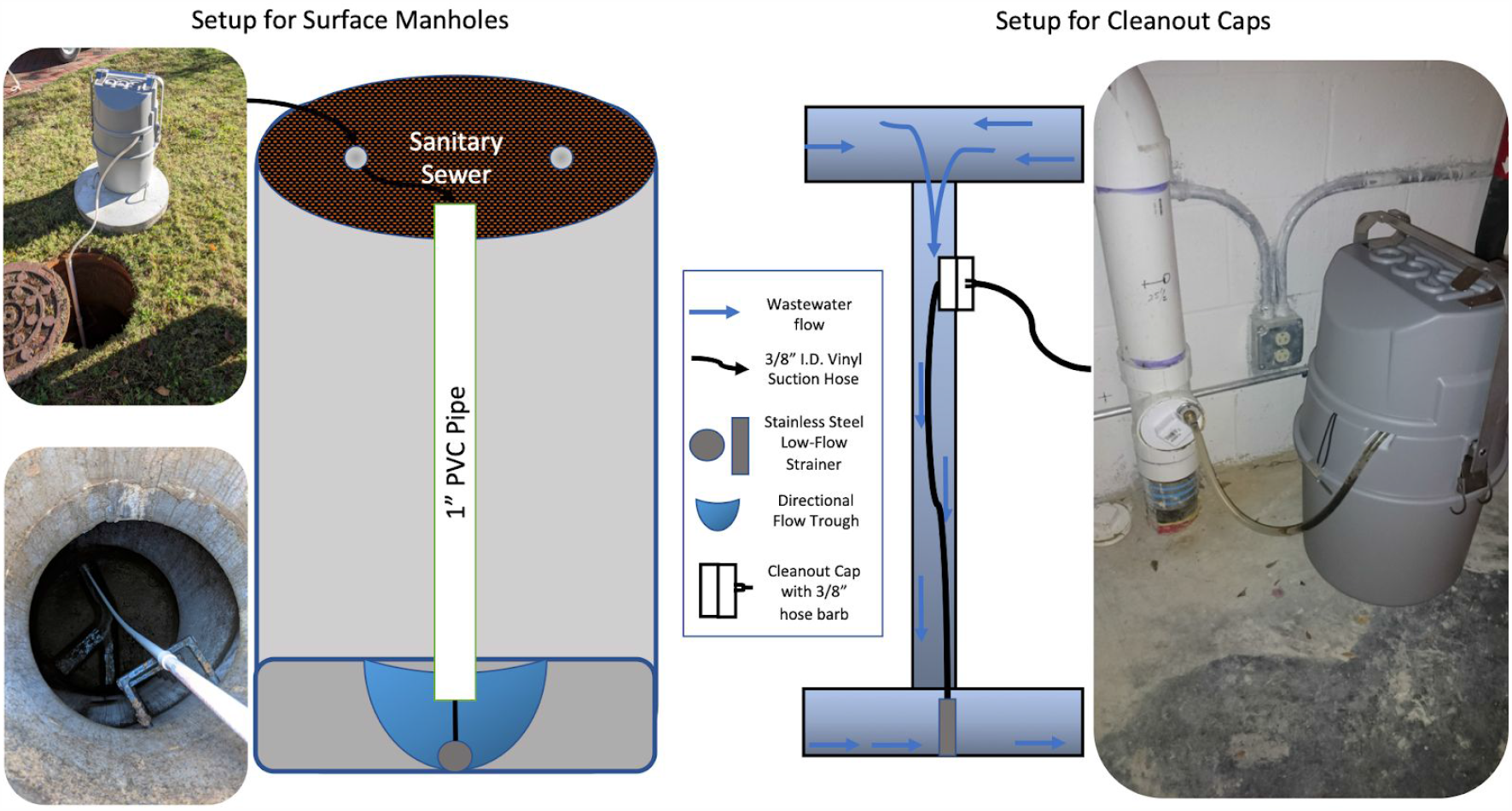
Autosampler setup for surface manholes and cleanout cap locations.

### Sample Concentration

Sample concentration is crucial for low titered virus to be detectable in RT-qPCR. There are several common methods used for concentration by other groups measuring the concentration of SARS-CoV-2 in wastewater, including PEG precipitation [17,18], centrifugation and ultrafiltration [19–21], direct extraction [22,23], and electronegative filtration [24,25]. In order to expedite project setup, we chose to use an established electronegative filtration protocol [24,26] that was already in use in the Munir lab in the context of a municipal wastewater project with the North Carolina Policy Collaboratory, now described in Ciesielski et al. 2020 [27]. Wastewater samples are concentrated following the method in [24,26] with a slight modification. A 40 mL aliquot is taken from each sample. pH of the aliquot is adjusted to 3.5 - 4.0 using concentrated HCl, followed by addition of 100X MgCl2 hexahydrate (2.5M) at a ratio of 1:100. As a process control, a known concentration of Bovine Coronavirus modified live vaccine (16445-3 Bovilis Coronavirus Calf Vaccine) is spiked into the composite sample at a ratio of 1:10000. Then the wastewater sample is passed through a 0.45 μm pore size, 47mm diameter, electronegative filter (HA, Millipore) via a disposable filter funnel (Pall Corporation) using a vacuum filtration manifold. 40 ml of PBS buffer is subjected to the same treatment as a filtration method control. In cases where filters can not be processed immediately, they are stored at −80°C.

### RNA Extraction

RNA extraction from environmental samples is typically complicated by the presence of inhibitors in the sample. The purity, concentration, integrity and cross-contamination jeopardies of the extracted RNA should be considered and mitigated, to preserve the quality of viral RNA for RT-qPCR analysis. Prior studies have used commercially available RNA extraction kits such as the RNeasy PowerMicrobiome Kit, the QIAamp Viral RNA mini kit, the NucleoSpin RNA virus kit, and the Zymo Research Viral RNA Kit for SARS-CoV-2 RNA extraction from wastewater [13,28–30]. The recovery of RNA and the performance of kits differs significantly based on the extraction kit used. Based on prior reports, supply chain considerations, and vendor information we settled on the QIAamp Viral RNA mini kit (Qiagen) for the process of RNA extraction. AVL buffer with carrier RNA (Qiagen) was used for extraction, as well as to suspend the filters prior to RNA extraction, as lysis buffers are known to stabilize viral material <u>[31]</u> and the carrier RNA can improve detection in low-concentration samples.

The filter collected in the concentration step is placed in a 2 ml microcentrifuge tube containing 1 mL AVL lysis buffer (Qiagen) and vigorously vortexed and incubated for 10 minutes to facilitate recovery of virus from the filter surface. RNA is then extracted from a 200 µL aliquot of the sample using the QIAamp viral mini kit (Qiagen) according to the manufacturer’s instructions. RNA is eluted with 60 µL of AVE buffer.

For infrequent cases where insufficient sample volume is collected for processing using the HA filtration/extraction approach, we instead use a direct extraction method, in which 1 mL of wastewater sample is mixed with 1 mL of Lysis buffer (Easymag, Biomerieux). 200 µL aliquots of the resulting mixture are then used in the extraction process.

### Molecular Detection of SARS-CoV-2

Reverse Transcriptase real time PCR (RT-qPCR) is still considered as a gold standard for the detection and quantification of pathogens [18,32]. Current alternative methods that have been explored for SARS-CoV-2 detection from environmental samples during the pandemic include droplet digital PCR (ddPCR) [33,34], third-generation sequencing [35,36], and Loop-Mediated Isothermal Amplification (LAMP) [37]. While we are equipped for all of these methods, RT-qPCR was the most immediately accessible starting point and has minimal bioinformatics needs, which was critical while most of our attention was focused on optimizing wet-lab operations.

Primer/probe sets have already been published for SARS-CoV-2 detection and quantification [38,39]. During the time frame discussed in this manuscript, we have relied on the CDC N1 primer alone for SARS-CoV-2 detection. In previous studies of influent wastewater, the CDC N1 based assay was found to be highly sensitive as compared to other available assays for detection of SARS-CoV-2. In a review of available publications and preprints describing wastewater based epidemiology protocols, Michael-Kordatou et al. [40] observed that the CDC N1 assay performed well for the detection of SARS-CoV-2 in wastewater samples in multiple studies.

We follow a commonly used RT-qPCR protocol to detect the presence of SARS-CoV-2 RNA in wastewater samples [24,25]. A one-step quantitative reverse transcriptase-polymerase chain reaction (qRT-PCR) is used to quantify SARS-CoV-2 RNA using the CDC N1 (Nucleocapsid) primer and probe set, 2019-nCoV CDC RUO Kit (IDT#10006713). All amplification reactions are carried out in a 20 µL reaction volume. The SARS-CoV-2 assay reaction includes 10 µL iTaq universal probes reaction mix (Bio-Rad, Hercules, CA), 0.5 µL iScript reverse transcriptase (Bio-Rad), 500 nM primers, 125 nM probe (IDT), and 5.0 µL of the extracted RNA template. The instrument used is the CFX96 qPCR thermocycler (Bio-Rad, Hercules, CA). Control plasmid containing the complete nucleocapsid gene from 2019-nCoV (IDT cat.#10006625) and nuclease free water are used as positive and negative controls, respectively. All samples are run in triplicate, and for each sampling event a series of three positive and negative controls is included in the plate.

For quality control purposes, both the extraction blank and reagent blank are amplified in a batch with samples to identify any carryover contamination. RNA extraction and RT-qPCR analysis are performed in separate rooms to prevent cross-contamination. Environmental samples often have inhibitors that can inhibit the amplification reaction. Inhibition is detected by running the diluted RNA template along with the main sample to observe the effect on Cq value [12]. For genome copy quantification, a standard curve for the N1 primer/probe set was generated using a series of tenfold serial dilutions of the positive control, with concentrations ranging from 100000 to 10 copies per reaction. The resulting best fit curve had an R^2^ value of 0.987, with slope and intercept of −3.203, and 38.987 respectively. Calculated primer efficiency under these conditions was 105.2%, which meets the requirement to be an efficient primer (usually 90-130%)[41].

Bovine coronavirus is quantified according to the published protocol by Decaro et al. [42], Primer and probe sequences are shown in Table 1. The BCoV RT-qPCR assay contains 10µL iTaq universal probes reaction mix (Bio-Rad), 0.5 µL iScript reverse transcriptase (Bio-Rad), 600 nM forward primer, 600 nM reverse primer, 200 nM probe, and 5.0 µL extracted RNA.

**Table 1:**
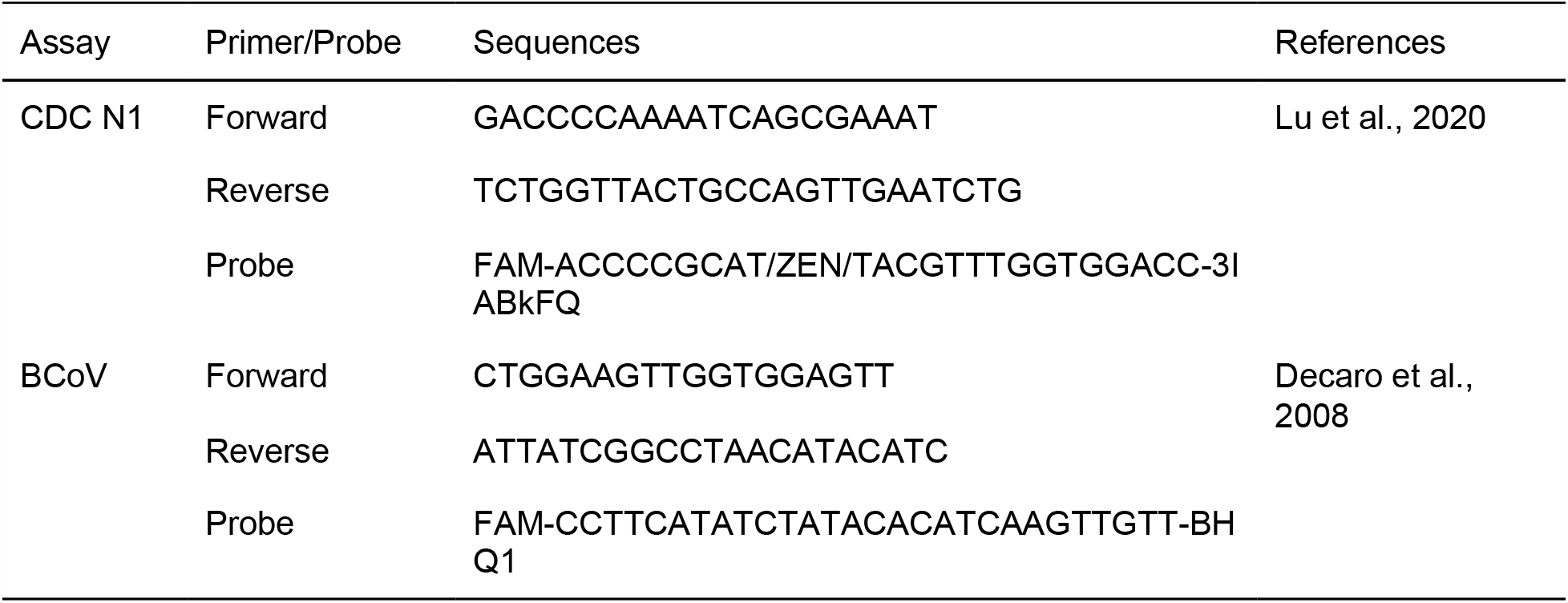
Primers and probe used in this study

The maximum cycle value for positivity of wastewater RNA samples is set at 45.0, although typical values of Cq in wastewater positives that have been validated by surge testing range from 33-37, which is within 2-4 Cq of the positive standard’s Cq value. For the purpose of administrative decision making, samples below the maximum Cq threshold are considered positive if SARS-CoV-2 is detected in all three technical replicates. A sample is otherwise marked as “suspicious” if only one or two replicates are positive.

## Results

### Typical Results for qPCR Detection of SARS-CoV-2

Typical results for an individual sampling event are shown in Table 2. Samples that were classified as positive had SARS-CoV-2 detected in all three sample replicates with mean Cq below 45. The high-Cq, positive signal reported in Building 13 resulted in RT-qPCR being repeated to confirm the positive due to the presence of a missing replicate, and then in the decision to surge test. Surge testing identified a single positive-testing individual among 177 tests performed.

**Table 2:** Typical daily results for detection of SARS-CoV-2 in wastewater samples. Samples collected from different locations at UNC Charlotte on 10-28-2020. Buildings are not identified by name, in keeping with university policy on COVID-19 reporting.

| Building | Sample Collected? | Volume | qPCR Amplification (Cq) Values |  |  | Mean Cq | Standard Deviation of Cq | SARS-CoV-2 Copies per Liter | Sample Status |
| --- | --- | --- | --- | --- | --- | --- | --- | --- | --- |
| Building 1 | Yes | 1.0 L | NA | NA | NA | NA | NA | NA | Negative |
| Building 10 | Yes | 3.0 L | NA | NA | NA | NA | NA | NA | Negative |
| Building 12 | Yes | 3.0 L | NA | NA | NA | NA | NA | NA | Negative |
| Building 15 | Yes | 3.0 L | NA | NA | NA | NA | NA | NA | Negative |
| Building 3 | Yes | 1.0 L | NA | NA | NA | NA | NA | NA | Negative |
| Building 4 | Yes | 1.0 L | NA | NA | NA | NA | NA | NA | Negative |
| Building 5 | Yes | 3.0 L | NA | NA | NA | NA | NA | NA | Negative |
| Building 6 | Yes | 3.0 L | NA | NA | NA | NA | NA | NA | Negative |
| Building 8 | Yes | 2.0 L | NA | NA | NA | NA | NA | NA | Negative |
| Building 9 | Yes | 3.0 L | NA | NA | NA | NA | NA | NA | Negative |
| Group 1 | Yes | 3.5 L | NA | NA | NA | NA | NA | NA | Negative |
| Quarantine | Yes | 3.5 L | NA | NA | NA | NA | NA | NA | Negative |
| **Building 13** | **Yes** | **3.5 L** | **37.12** | **36.63** | **NA** | **36.38** | **1.05** | **9.77E+03** | **Positive** |
| **Group 2** | **Yes** | **1.0 L** | **34.96** | **33.91** | **31.12** | **33.33** | **1.98** | **8.95E+04** | **Positive** |
| Building 11 | No | 0 | NA | NA | NA | NA | NA | NA | Unknown |
| Building 14 | No | 0 | NA | NA | NA | NA | NA | NA | Unknown |
| Building 2 | No | 0 | NA | NA | NA | NA | NA | NA | Unknown |
| Building 7 | No | 0 | NA | NA | NA | NA | NA | NA | Unknown |
| Isolation | No | 0 | NA | NA | NA | NA | NA | NA | Unknown |

### Prevalence of Wastewater Positives During Fall 2020

During the period from September 28, 2020 to November 23, 2020, the UNC Charlotte wastewater surveillance team collected and processed 332 out of an expected 475 wastewater samples from 19 building sites. Failure to collect samples generally occurred due to low flow at collection sites or problems with placement of autosampler intakes, and these issues were gradually corrected over the course of the semester. Of the samples collected, 40 were classified as positives, and 5 others were labeled suspicious due to positive signal appearing in only one qPCR replicate. 16 of these positives were “new” positives, defined as the first positive wastewater test in a given building in greater than 10 days. 8 of these resulted in a wastewater surge test. 19 additional samples were positive due to the sampling of the isolation or quarantine buildings. Figure 4 summarizes the sample collection and testing results.

**Figure 4:**
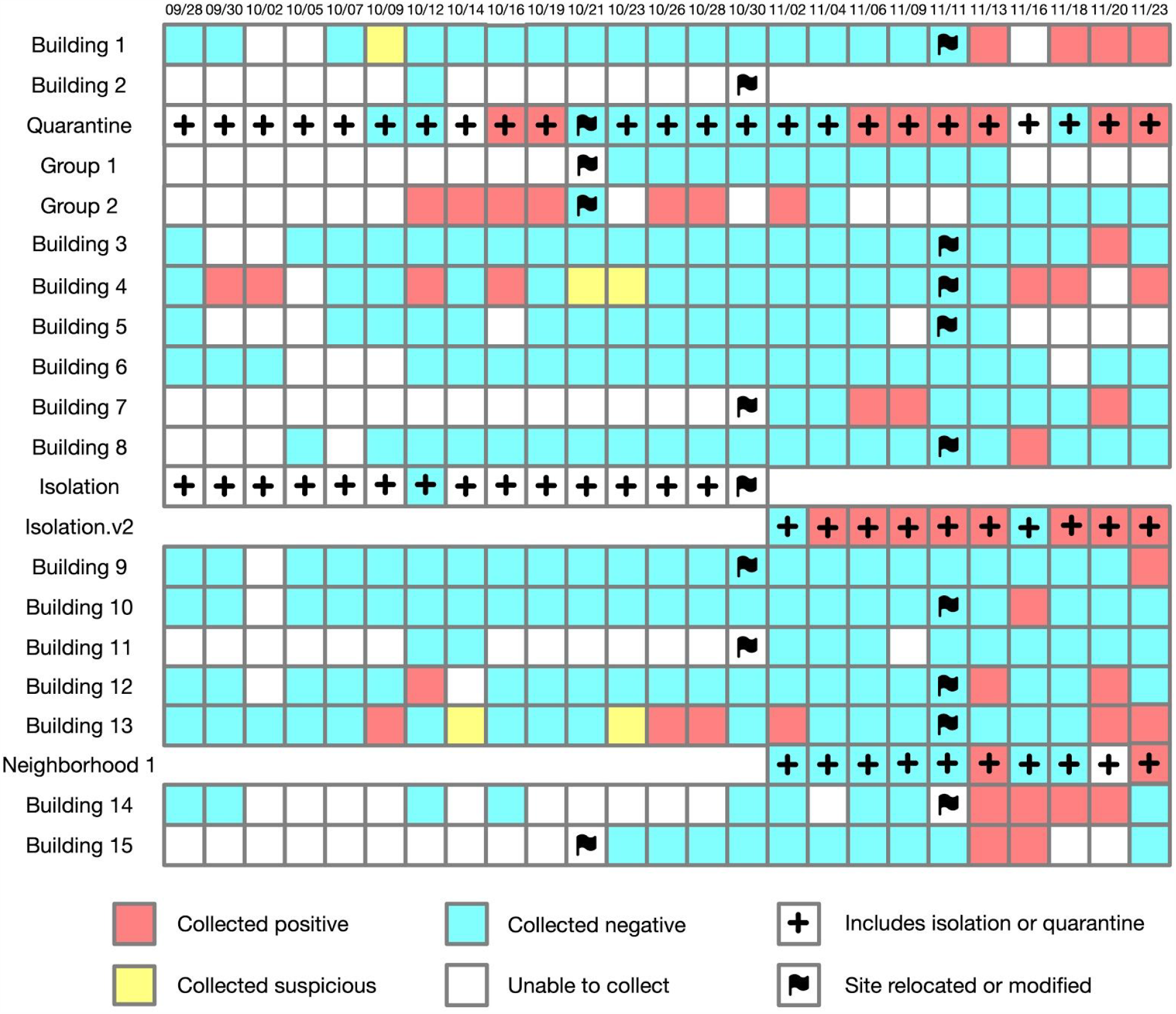
Overview of sample collection and detection outcomes over the 8-week on-campus session.

Over the course of the 8 week in-person semester, the number of observed positive wastewater samples at each sampling event gradually increased. From a purely procedural point of view, this can be partly explained by improvement in our ability to collect samples at some sites by relocation or reconstruction of the site. But within the larger context of a gradual increase in positivity rates in Mecklenburg County, NC, the increase in cases on campus was also to be expected. The number of new daily positive cases in Mecklenburg County as a whole [43] and the number of buildings detected as positive on each sampling event are correlated over this time frame (Figure 5).

**Figure 5:**
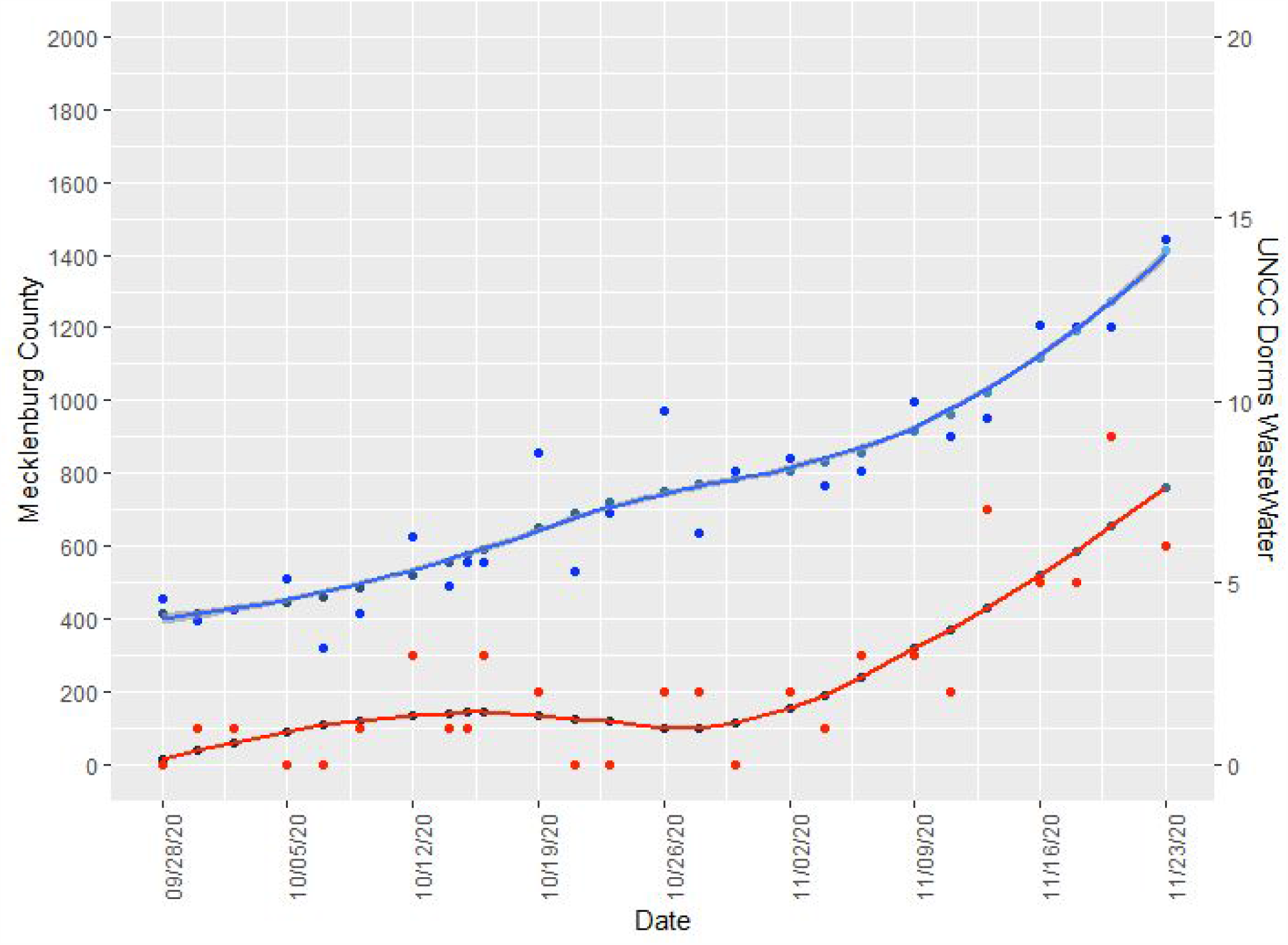
Total number of SARS-CoV-2 positive wastewater samples from UNCC dormitories (red, right axis) and daily new positive cases in Mecklenburg county (blue, left axis) from September 28 to November 23, 2020. Dots represent the original data values while the blue and red curves are fit to the data using Loess Smoothing. Pearson correlation coefficient between the two series is 0.769.

### Wastewater-triggered Surge Testing Events and Outcomes

When we began the wastewater surveillance project, University of Arizona had just reported an early success with isolation of COVID-19 positive students following detection of the virus in wastewater [8]. However, we had little idea of how soon to expect actionable positive results on our own campus. On September 24 and 25, students began arriving from all over North Carolina and beyond. North Carolina had experienced a consistent level of coronavirus infection all summer without a dramatic spike in case numbers [44]. September 30, our second day of sampling brought the first of many wastewater positives, while the campus as a whole was still largely free of known COVID-19 cases. Because of a lack of any prior information suggesting COVID-19 spread in the affected building, the entire student population of that building was asked to shelter in place and tested the following day. This event gave the university an opportunity to shape its strategy for responding to future events (Figure 6).

**Figure 6:**
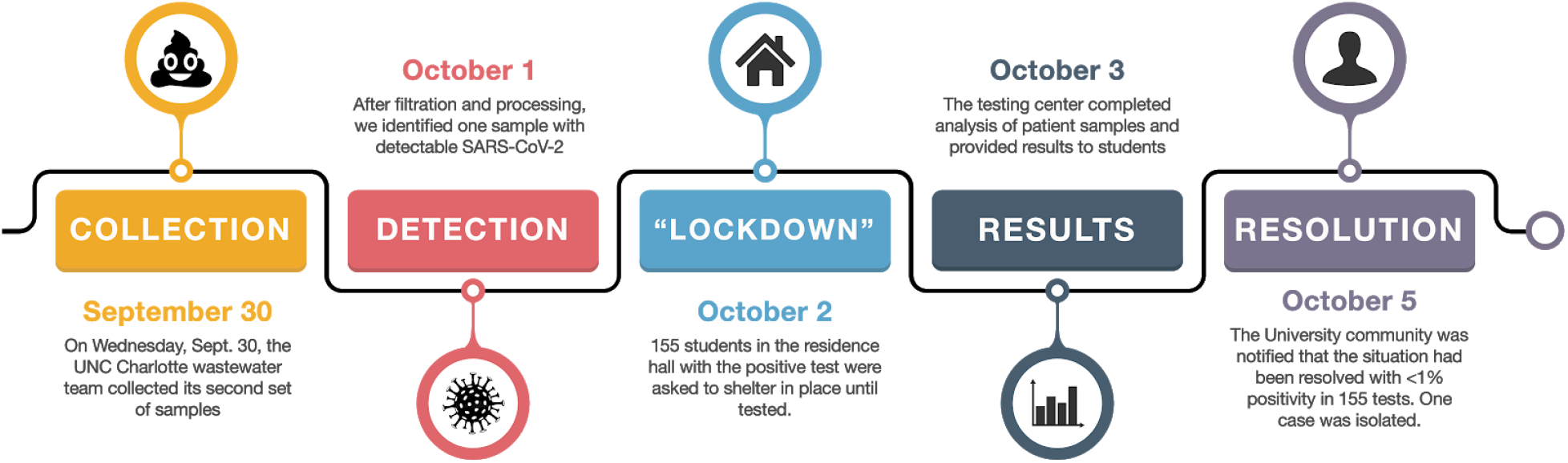
Timeline of campus actions after the first positive wastewater event on the UNC Charlotte campus, Sept. 30 2020.

The next two weeks were a relatively quiet period in terms of COVID-19 cases on campus. In mid-October, a cluster was detected via contact tracing, in the absence of any wastewater signal. This alerted us to the need to relocate some of our initial sampling sites to provide more complete coverage of large buildings. We began an analysis of plumbing and architectural plans that resulted in selection of new sampling sites. As the semester progressed, wastewater-triggered events became more frequent, both due to improved ability to collect samples and to increasing prevalence of COVID-19 in the community as a whole. In the week leading up to student move-out prior to the Thanksgiving holiday, four buildings had to be surge tested consecutively.

On only one occasion during this timeframe did a wastewater-triggered surge testing effort fail to identify any positive case. On November 4, three buildings (smaller buildings sampled as a pool at one manhole) were selected to be tested based on an ongoing positive wastewater signal. However, only 70% of students identified as living in those buildings complied with the initial testing request, and the source of the positive signal was not immediately identified.

Table 3 shows outcomes of the eight wastewater-triggered surge tests for COVID-19 at UNC Charlotte during the in-person portion of fall semester 2020, Sept. 28-Nov. 23. Reporting of these outcomes to the public by the Office of Emergency Management has changed over the course of the semester. On Nov. 4, the university switched from reporting number tested to reporting percentage of the building tested. On Nov. 18, the university reported the results of three separate surge tests at the same time with two of the buildings grouped as a cluster based on contact evidence. We are currently working with administration groups to develop a more consistent approach for retaining and describing surge test data and metadata internally. Overall, when all University notices reporting cases and clusters in the community during fall semester were analyzed, clusters detected in surge testing events triggered by wastewater surveillance were consistently smaller in size than clusters detected by other methods. (Figure 7). While this is based on only the small amount of publicly available data for fall semester, it is an encouraging trend in that context. A t-test between the two groups finds that the difference is significant with a p-value of 0.000546.

**Table 3:** Outcomes of wastewater-triggered surge tests during fall 2020.

| NinerNotice Date | Positive WW Sample Collected | Site ID | Site Cq | Number Tested | Positivity Rate Reported |
| --- | --- | --- | --- | --- | --- |
| Oct. 2 | Sept. 30 | Building 4 | 42.3 | 155 | <1% |
| Oct. 28 | Oct. 26 | Building 13 | 39.0 | 176 | <1% |
| Nov. 4 | Nov. 2 | Group 3 | 35.8 | 70% | 0% |
| Nov. 11 | Nov. 9 | Building 7 | 33.8 | 85% | <1% |
| Nov. 16 | Nov. 13 | Building 14 | 36.2 | 82% | <2% |
| Nov. 18 | Nov. 13,<br>Nov. 16,<br>Nov. 16, | Building 1,<br>Building 4,<br>Building 10 | 35.7,<br>30.6,<br>31.6 | 86%,<br>77%,<br>92% | <1%,<br><3%,<br><3% |

**Figure 7:**
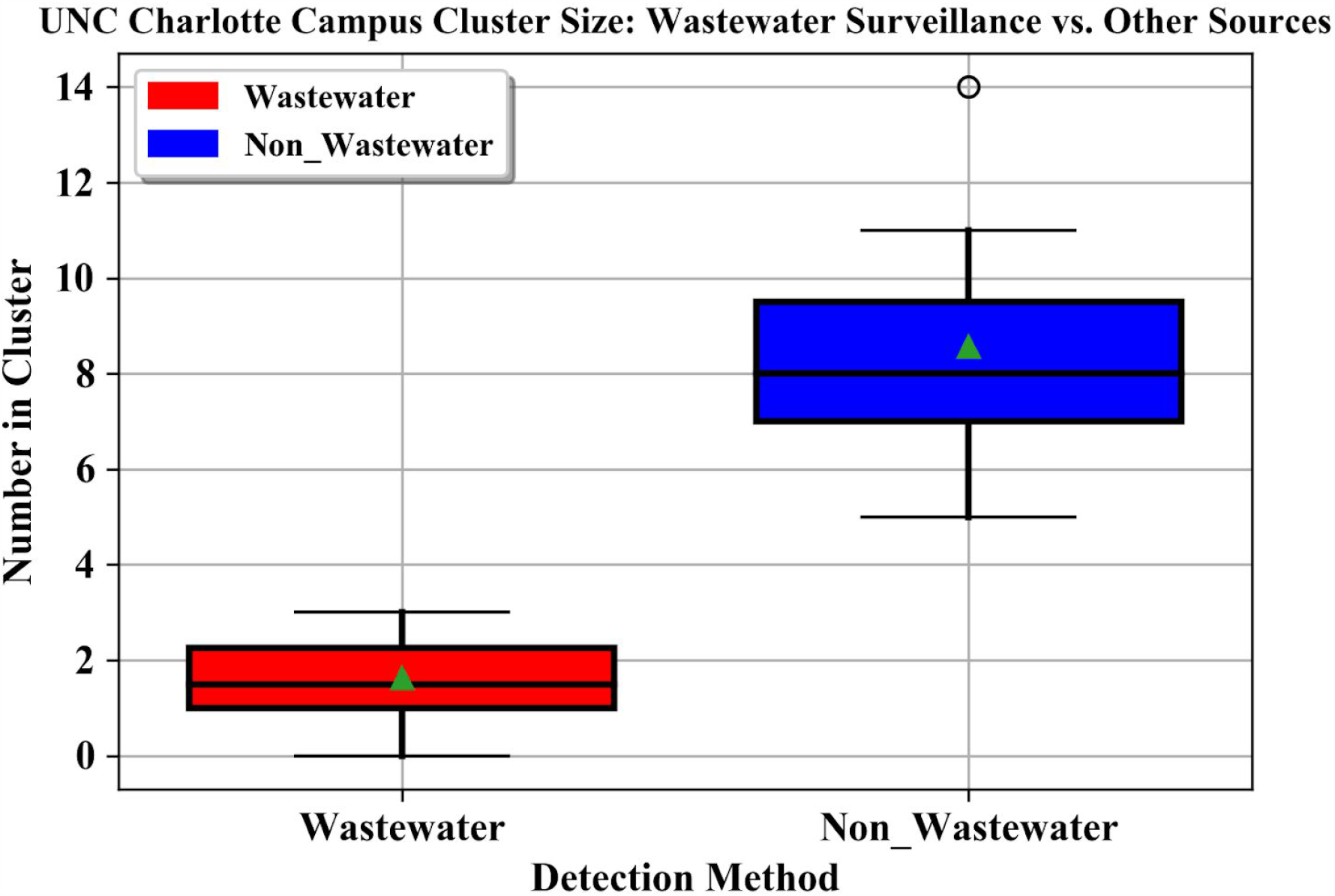
Number of positive cases detected in wastewater-triggered surge testing events vs. number of cases in other reported campus clusters during the fall semester (Sept. 1-Nov. 23, 2020). This includes one cluster event dated prior to the start of wastewater sampling, which involved students who had already moved into dorms for the original Sept. 7 start date. Numbers are extracted from publicly available reports (Niner Notices, [45]) made available to the University community and may not include students who reported late for required surge testing or cases that evaded surge testing.

In addition to detecting presymptomatic cases, the wastewater program has been able to provide corroborative evidence for the existence of cases suggested by contact tracing, and resumption of negative wastewater signal in a building has also been interpreted as an indication that an outbreak in a building has been successfully contained.

## Discussion and Conclusions

In fall of 2020, UNC Charlotte implemented a wastewater testing program that provided actionable results for University administrators. The program will be continued in the first half of 2021, and potentially beyond depending on how quickly we progress towards the resolution of the pandemic. The break between semesters provides an opportunity to evaluate and change various aspects of the wastewater surveillance program. Returning to the six key methodological choices we outlined above (where, when, and what type of sample, concentration method, extraction method, and detection method) we found that we had landed on acceptable protocols in some cases, while in other cases further optimization is needed.

The move to sample entire buildings from exterior manholes provided the best coverage of the campus, and dilution by other sources did not negatively impact our ability to identify individual COVID-19 cases within building populations. When it is possible to use a single site to sample an entire building, this is adequate for the purposes of the program at current occupancy levels. In the few cases where plumbing architecture requires us to split a building into zones, the University administration still prefers to surge test the entire building based on a signal within one of the zones, and so splitting additional buildings into zones is not considered necessary at this time. To inform sewershed modeling, we plan to add a group of “neighborhood” sites that capture larger areas of the campus, especially those buildings that are frequented by essential staff who must work in person, and academic buildings that also house active research laboratories. We will also sample several non-residential buildings that are known to have experienced relatively high student traffic during the fall semester, such as the library, the student union, and the university recreation center. While data from these sites will not be directly actionable, in that the university administration can not specifically identify who may have passed through those buildings, the neighborhood sites and high-traffic buildings will provide an understanding of background levels of SARS-CoV-2 on the campus as a whole. A thrice-weekly sampling schedule is adequate for the University’s purposes, and both the wastewater surveillance team and the larger Testing, Tracing and Monitoring Task Force agreed that there is no apparent need to test wastewater at a higher frequency. However, shortening the time between the start of autosampling and reporting of results is considered a priority. Currently, autosamplers are started at approximately 8 a.m. on day one of the lab’s three-day working cycle, and results are reported to the administration by 2 p.m. on day three. Surge testing of students requires preparation of personnel and materials, so that the earliest a round of testing will start after a positive report is generally the following morning. This can result in a positive signal from one case overlapping into a second and even third cycle of sampling, depending on how long it takes to test the affected areas and move infected students to isolation. There are two “pain points” in the current protocol that have the potential to shorten this timeframe -- first, the length of the autosampling window, and second, the use of the well-validated but time-consuming HA filtration process for the concentration step. Optimizations for both of these steps are in progress. Testing the downstream impact of a sampling window change on workflow outcomes is a high priority for the pre-semester period in January. We are also planning to replace the lengthy HA filtration step with a faster concentration protocol so that results can be reported by the end of the day the sample is collected, rather than waiting into a second day, and have begun a side-by-side comparison of the two protocols.

While we are generally satisfied with the performance of our extraction and detection steps for routine sample processing, continuing optimization of detection protocols is also a priority for the project. While use of N1 as the sole indicator of presence of SARS-CoV-2 is supported by the literature, the BioRad CFX96 platform permits multiplexing of N1 and N2 primer/probe sets, or multiplexing with components of other probe sets in use worldwide. As we look to generalize the building wastewater monitoring strategy to other scenarios, it is also of interest to multiplex SARS-CoV-2 primer/probe sets with process controls and with diagnostic primer/probe sets for other infectious diseases. Availability of low-cost sequencing also holds the promise of identifying individual strains which are spreading in the campus environment, and in connecting the history of wastewater positives with specific patient tests.

Finally, the university’s mix of testing, self-reporting, contact tracing and wastewater sampling has created a wealth of potentially interesting metadata. Once remaining issues of student privacy and data access are resolved, these data will support modeling of the relationship of wastewater signal to COVID-19 prevalence on campus and in the surrounding region. Integration of wastewater results with building-level aggregate reporting of positive tests, close contacts and voluntary symptom reports will enable development of a wastewater dashboard for internal administrative use. We will test analytic approaches to determine if the implied decision tree used by administrators in choosing when to surge test can automatically make the same recommendations, and if not, what an accurate formal representation of that decision process would be (Figure 8).

**Figure 8:**
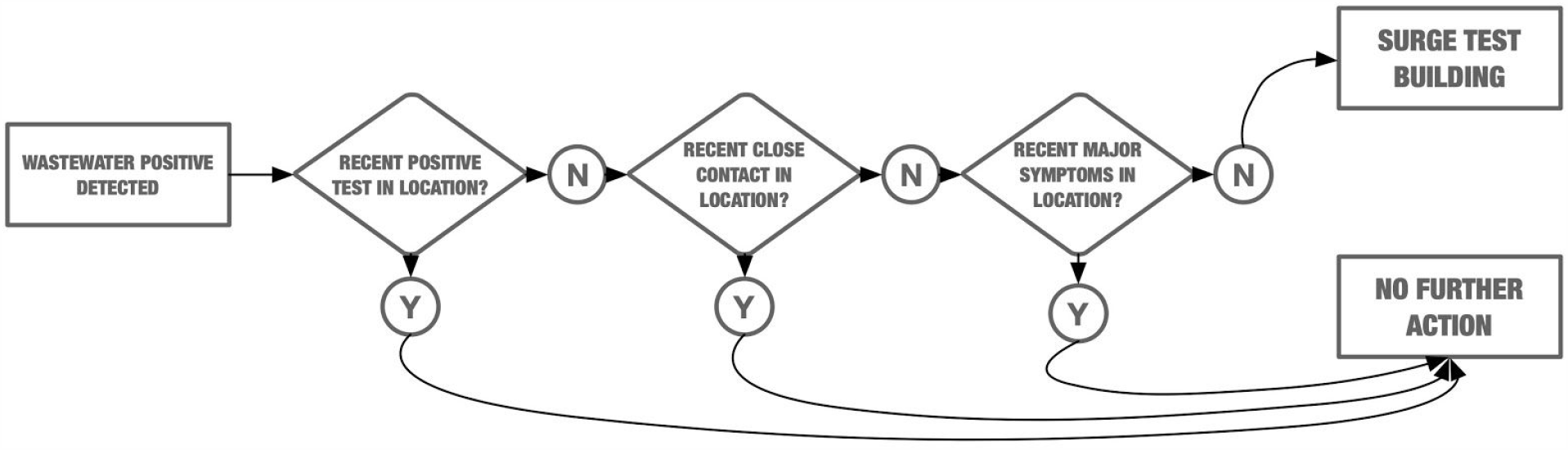
Decision tree representation of the process used by UNC Charlotte administrative group when deciding whether or not to follow up positive wastewater signal with surge testing. Based on conversations within the UNC Charlotte Testing, Tracing and Monitoring Task Force during Fall 2020.

With a vaccine on the horizon, the future of a COVID-19 specific wastewater monitoring program is likely not long-term, but its lifespan depends on how quickly vaccines are rolled out to the general population and adopted. National policy leaders have recently gone on record with statements [46] that vaccination rates above 75-80% will ultimately be needed to bring the COVID-19 pandemic to an end. It is predicted that even with gradual introduction of vaccines, some types of reopening policies may result in continued waves of outbreaks [47], and that vaccine hesitancy [48] may be a significant challenge in achieving sufficient vaccine coverage in the short term. As students and employees are vaccinated, it will be of great interest to follow the wastewater and other evidence streams on campus to detect when vaccines begin making an impact on spread, and at what point a resumption of in-person operations at normal density is safe. Another factor in the timeline of the COVID-19 pandemic is the emergence of new strains such as B.1.1.7 [49], their ability to evade antibodies elicited by current vaccines, and the ability of labs to adapt to detection and monitoring of multiple strains. However, the lessons learned from COVID-19 are applicable to monitoring of other pandemic diseases and potentially also to management of recurring seasonal outbreaks such as influenza, which may not be as urgently threatening as COVID-19, but still impact community health and operations.

It is important for those considering implementing wastewater surveillance on college campuses and in other group living situations to consider the context in which positive pathogen signals in wastewater will be evaluated, and especially how wastewater information will be combined with other information to decide when a positive result is actionable. Without insights from symptomatic testing, contact tracing and voluntary symptom reporting, positive wastewater samples would likely have triggered many more costly surge testing events at UNC Charlotte. As part of a comprehensive approach to pandemic mitigation, wastewater surveillance is a valuable tool for identifying undetected new or presymptomatic COVID-19 cases in dormitories. While time will tell whether the WBE approach is as useful in future pandemic scenarios with different viral transmission patterns, it is clear that this approach is worth refining. Development of wastewater surveillance infrastructure plans, analytics and decision-support tools for organizations using WBE to protect residents and workers will improve readiness for the next pandemic. The approach is also likely to be useful in other high-density and congregate living situations, such as apartment complexes, long term care facilities, and military facilities, and facilities with a well defined population of users, such as schools and corporate campuses.

## Data Availability

Data and protocols are available from the authors upon request.

## Acknowledgements

The UNC Charlotte wastewater group would like to thank Chancellor Sharon Gaber, Provost Joan Lorden, and Richard Tankersley, Vice Chancellor for Research and Economic Development and his team for strong institutional support of this project, Lawrence Mays, Chair of the Department of Bioinformatics and Genomics, and John Daniels, Chair of the Department of Civil and Environmental Engineering, for facilitating setup of the project in their research spaces, Rachel Noble and colleagues in the North Carolina Policy Collaboratory for early research into wastewater sample processing protocols that was leveraged in this project, Greg Cole and his team in Facilities Management for construction and plumbing support, and the Testing, Tracing and Monitoring Task Force (Robert Jones, Medical Director, Keith Carnes, Angelica Martins, Emily Stewart, Patrick Versace, and Susan Messina) for ongoing discussion of the institutional COVID-19 management strategy at UNC Charlotte.

## Individual Contributions

CG -- developed the project concept and overall strategy, coordinated wastewater surveillance and reporting with other components of the campus COVID-19 strategy, analyzed data, drafted the manuscript

KL -- designed and implemented the field sampling operation, drafted the manuscript

NM -- implemented and managed the sample processing strategy, trained and managed the sample processing team, maintained and analyzed the data, drafted the manuscript

MAIJ -- optimized qPCR protocols, analyzed the data, drafted the manuscript

VBB -- established methods for HA filtration and extraction, trained and managed the sample processing team, maintained and analyzed data, drafted the manuscript

LRB, KH, JL, NS, IY, CQ, MR, JK, BN -- collected, processed and analyzed samples, optimized lab and field protocols

WT -- prototyped data analysis and visualization infrastructure to support automated reporting

SA -- analyzed campus sewershed to optimize future sampling

DC -- analyzed plumbing and architectural plans to define sampled zones

JS -- developed the project concept and overall strategy, coordinated laboratory operations, equipment acquisition, biosafety protocols and training, drafted the manuscript

MM -- developed the project concept and overall strategy, established and optimized overall lab workflow for sample processing, extraction and qPCR detection, drafted the manuscript

